# Postnatal depression, to treat or not to treat: Long-term consequences of postnatal selective SSRI treatment on mother and child

**DOI:** 10.1101/2022.05.31.22275818

**Authors:** Chao-Yu Liu, Eivind Ystrom, Tom A. McAdams

**Affiliations:** Social, Genetic and Developmental Psychiatry Centre, Institute of Psychiatry, Psychology and Neuroscience, King’s College, London, UK; PROMENTA Research Center, Department of Psychology, University of Oslo, Oslo, Norway; Department Mental Disorders, Norwegian Institute of Public Health, Oslo, Norway; Pharmaco-Epidemiology and Drug Safety Research Group, School of Pharmacy, University of Oslo, Oslo, Norway

**Author notes:** Address for correspondence: Dr Chaoyu Liu, Social, Genetic and Developmental Psychiatry Centre, King’s College, 16 De Crespigny Park, London SE5 8AF. Social, Genetic and Developmental Psychiatry Centre, Institute of Psychiatry, Psychology and Neuroscience, King’s College, London, UK.

**Keywords:** postnatal depression, long-term effect, antidepressants, SSRI, externalizing behavior, development

## Abstract

**Background:** Although selective serotonin reuptake inhibitors (SSRIs) are recommended for postnatal depression (PND) treatment, a lack of evidence regarding the long-term consequences of postnatal SSRI use have resulted in treatment hesitancy.

**Methods:** The current study used longitudinal data on a total of 60654 mother-child dyads enrolled in the Norwegian Mother and Child Cohort Study (MoBa) to examine associations between PND and maternal outcomes (depression and anxiety, relationship satisfaction) and child outcomes (motor and language development, emotional and behavioral problems) from birth to postpartum year 5. We tested whether postnatal SSRI treatment moderated the effects of PND on maternal and child outcomes. A propensity score was used to control for pre- /antenatal factors that impacted the probability of receiving SSRIs.

**Results:** PND was associated with poor maternal mental health outcomes and problems in child development. Use of SSRIs mitigated the associations between PND and later maternal mental health distress and child externalizing behaviors up to postpartum year 5. We found no evidence to indicate that the use of SSRIs was associated with increased risk of negative outcomes in emotional, behavioural, motor and language development in offspring.

**Conclusions:** Our findings suggest that SSRI treatment for PND may bring benefits in the long term by attenuating the detrimental associations between PND and subsequent maternal depression and child externalizing behaviors. The study provides valuable information for clinicians and women with PND to help make informed decisions regarding treatment.

## Introduction

Postnatal depression (PND) is a common psychiatric disorder that affects 10 to 15% of women in the first year after childbirth [1]. Depression is among the most influential illnesses on the global burden of mortality and disability [2]. The clinical presentation of PND resembles that of major depressive disorder, which is characterized by impairing symptoms of depressed mood, lack of interest, fatigue, poor concentration, and suicidal thoughts [3]. In addition to experiencing depression during the immediate postpartum period, women affected by PND are more susceptible to recurrent depressive episodes in subsequent pregnancies and display higher levels of depression and anxiety in the years following childbirth [4, 5]. It is noteworthy that PND also predicts adverse outcomes in the wider family. PND is associated with long-term partner relationship problems [6] which, in turn, is a risk factor for perinatal depression and prolonged PND courses in affected women [7-9].

Children born to mothers with PND show increased difficulties in social adaptation, delayed cognitive development, and higher levels of behavioral and emotional problems [10, 11].

Moreover, persistent PND symptoms further the risk to child’s development, whilst remittance of PND is associated with normalization of behavioral problems and psychopathology of the affected children [12, 13]. Such findings suggest that effective intervention for PND may mitigate some of the negative consequences associated with the condition.

The effectiveness of antidepressants in the management of PND has been proven by meta-analysis of data from randomized control trials [14]. Selective serotonin reuptake inhibitors (SSRIs) are often the medication of choice to treat PND due to better safety profiles and tolerance compared to other antidepressants such as tricyclic antidepressants (TCA) and monoamine oxidase inhibitors (MAOi) [15]. However, there is insufficient evidence regarding the long-term consequences of postnatal SSRI use for women and their children [14]. Findings regarding associations between *ante*natal exposure to SSRIs and delay in motor and language development and increased emotional problems of children raise concerns about neurodevelopmental consequences associated with *post*natal SSRI exposure [16-19]. Although studies have pointed out that SSRIs recommended in the treatment for PND produce undetectable to very low serum levels in the exposed infants [20, 21], data on longitudinal neurodevelopmental effects are lacking [14]. This lack of evidence regarding long-term consequences for children has contributed to a lack of clarity and confidence regarding the appropriateness of pharmacological treatment for women affected with PND. This is important because inadequate treatment potentially increases the risks of poorly controlled PND symptoms and a prolonged disease course, resulting in more functional impairment and impaired mother-infant bonding [22].

In the current study we set out to examine the risks and benefits associated with postnatal SSRI use in the management of PND. Using data collected from pregnancy to postpartum year 5, we examined short- and long-term maternal mental health outcomes, alongside child developmental outcomes associated with PND. We then explored whether SSRI treatment moderated associations between PND and the study outcomes. It is noteworthy that several personal and environmental factors may influence treatment for PND, the use of SSRIs and associated outcomes. For example, women with more severe PND symptoms, prior lifetime depression diagnosis and prior experience with pharmacological intervention for depression are more likely to receive SSRIs for PND [22, 23]. Careful control for potential sources of confounding is therefore crucial to avoid pre-existing differences between treated and untreated women leading to incorrect conclusions regarding associations between SSRI use and maternal and child outcomes. To control for confounding factors that might partially or completely explain associations between PND and/or SSRI use and our outcomes, we included a propensity score derived from prenatal and antenatal variables that differentiated PND and/or treatment status as a covariate to adjust for probability of postnatal SSRI exposure.

Based on available data on infant’s response to maternal use of SSRI [24] and the positive outlook associated with well managed PND [25], we hypothesized that SSRI treatment can bring more benefits than harm in the long term for affected women, their children, and family.

## Methods

### Sample

Data were obtained from the Norwegian Mother and Child Cohort Study (MoBa), a prospective population-based cohort study in Norway. MoBa recruited women who attended routine ultrasound examination in week 17-18 of pregnancy from year 1999 to 2008. Women were invited to participate multiple times when they had more than one pregnancy. Among 277,702 invitations sent, over 95,000 women and 114,000 children were enrolled (participation rate of 41%) [26]. Participating families received questionnaires for child development and maternal health conditions across several assessments from gestational week 17 till present. Participant data can also be linked with the Medical Birth Registry of Norway (MBRN), which registers pregnancy outcomes in Norway after the 12^th^ gestational week. This study is based on version 12 of the quality-assured data release in January 2019. The current study included women with data on their depression symptoms at gestation week 30 and postpartum month 6, and data on self-reported medication use for mental health problems at postpartum month 6. To maximize our study sample, we included all pregnancies and randomly selected 1 child from each twin birth.

### Measurement

#### Prenatal and antenatal factors & potential confounding variables

Prenatal maternal factors included self-reported current and/or lifetime depression history, educational level and income at gestation week 17. Educational level was reported on a scale of 1 to 6, with 1 indicating ‘secondary school’ and 6 ‘higher education (university and above)’. Income was yearly gross income of the mothers including benefits and other allowance, from 1 as ‘no income’ to 7 as ‘over 500.000 NOK’.

Antenatal factors included alcohol and tobacco consumption during pregnancy, maternal age at childbirth, parity and maternal depression and anxiety. Antenatal alcohol consumption was collected at gestational week 30. We categorized the response to ‘ever use’ alcohol if the women reported alcohol consumption across any of the three trimesters during pregnancy. ‘Ever use’ tobacco was defined as tobacco smoking during the last three months of pregnancy reported at postpartum month 6.

Maternal depression and anxiety symptoms were assessed with the 8-item short version of the Hopkins Symptom Checklist (SCL-8) [27] at gestational week 30 and multiple time points after childbirth (postpartum month 6, years 1.5, 3 and 5). The SCL-8 captures symptoms of depression and anxiety with high correlation (r=0.94) with the original instrument.

Cronbach’s alpha was estimated at 0.88 in MoBa [28].

### Postnatal Depression (PND)

Self-report SCL-8 and the Edinburgh Postnatal Depression Scale (EPDS) [29] were used to measure women’s depression symptoms at postpartum month 6 (symptoms during the past week at the time of the questionnaire). Each item of the EPDS was rated on a Likert scale ranging from 0 (never) to 3 (most of the time). The 6-item version of the EPDS (EPDS-6) used in the MoBa study has been shown to correlate highly (r=0.961) with the 10-item full version. Cronbach’s alpha for the EPDS-6 was 0.84 in the MoBa sample. In the present study we defined women as experiencing postnatal depression (PND) with score ≥ 7 at the EPDS-6. This threshold has been validated in previous studies [30, 31]. The dichotomous PND status was coded separately for each pregnancy. The EPDS-6 and SCL-8 assessed at postpartum month 6 correlated at 0.70.

### SSRI-treated PND dyads and non-SSRI treated PND dyads

Mothers were asked to report any medications they had taken at postpartum month 6. The reported medication was identified using the Anatomic Therapeutic Classification (ATC) system of the World Health Organization. Because SSRIs are the most prescribed and researched antidepressants for perinatal mental health problems [14] and are recommended as the first-line treatment by multiple treatment guidelines [15, 32], the current study focused exclusively on the use of SSRIs. Mothers with PND who reported treatment with only anxiolytics (ATC class N05B), hypnotics (ATC class N05C) or non-SSRI antidepressants (ATC class N06AA, N06AF, N06AX) were excluded from analyses (n=78). If the mother identified as having PND reported SSRI (with ATC class N06AB) use, she was classified as a ‘SSRI-treated PND’. Conversely, if the mother did not report SSRI use, she was classified as a ‘non-SSRI-treated PND’. We excluded mothers who reported SSRI use but did not meet the threshold for PND (n=177).

Three mutually exclusive groups of mother-child dyads were identified: non-PND (n=52038), non-SSRI-treated PND (n=8440) and SSRI-treated PND (n=176). Due to the small number of participants who reported SSRI use, analyses comparing individual SSRIs were not performed.

### Maternal outcomes

Maternal depression and anxiety were assessed with the SCL-8 at postpartum years 1.5, 3 and 5. Partner relationship satisfaction was assessed via maternal report on the 5-item short version Relationship Satisfaction Scale (RSS) at postpartum month 6, and years 1.5 and 3. Items were rated on a 6-point scale to reflect relationship satisfaction (e.g. ‘I am very happy with our relationship’) and partner relationship quality (e.g. ‘my partner is generally understanding’). The RSS demonstrates good psychometric properties [33] and shows high predictive validity of overall life satisfaction [34].

### Child outcomes

Child outcomes comprised maternal reports on child internalizing and externalizing behaviors, ADHD symptoms and motor and language development. Internalizing and externalizing behaviors were measured with selected items from the Child Behavior Checklist (CBCL) [35] at ages 1.5, 3 and 5 years. There were in total 3 items for internalizing behaviors and 8 items for externalizing behaviors overlapping across the three time points. Composite scores for internalizing and externalizing behaviors were calculated separately at each time point.

Higher scores suggested more severe problems.

ADHD symptoms were assessed with items from the Conners Parent Rating Scale-Revised, Short Form (CPRS-R) [36] when the child was 5 years old. We calculated a composite score from the 12 available items of the CPRS-R tapping inattention and hyperactivity/impulsivity in the MoBA questionnaire to indicate ADHD symptom level.

Motor and language development were measured with items from the Ages and Stages Questionnaire (ASQ) [37] when the child was 1.5 and 3 years old. The Norwegian version of the ASQ has shown high agreement (84%) with standardized assessment tests and effective predictive validity for children with developmental delay [38]. Composite scores for motor and language development were calculated for ages 1.5 and 3 years respectively, with higher scores suggesting better performance.

## Data Analysis

Analyses were conducted with R (R version 4.0.3)[39]. We used the False Discovery Rate (FDR) approach at FDR<5% to adjust for multiple testing [40].

### Prenatal and antenatal factors associated with PND status and SSRI treatment: creating a propensity score to adjust for potential confounding

We compared prenatal and antenatal factors between non-PND and PND dyads and between SSRI-treated- and non-SSRI-treated PND dyads. Univariable and multiple logistic regressions were used to identify prenatal and antenatal factors differentiating PND status and SSRI use. Factors that differentiated SSRI use in multiple regression were treated as potential sources of confounding and were used to generate a propensity score to adjust for SSRI exposure probability for each pregnancy. This propensity score was included in the following analyses as a covariate to control for potential confounding in the associations between PND, SSRI use and study outcomes.

### Effects of PND on maternal and child outcomes and the moderation of SSRI

To study associations between postnatal depression and maternal and child outcomes, we examined associations between the global score of the SCL-8 at postpartum month 6 and study outcomes. We chose the SCL-8 because it was repeatedly measured in the follow-up period to reflect women mental health while the EPDS score was used to categorize PND status. Multilevel models with random intercepts (R package ‘lcmm’) [41] at the maternal level were applied to account for non-independence in our study sample (multiple pregnancies nested within mothers). The models were then expanded to include covariates and the interaction between SSRI and SCL-8 to examine whether receiving SSRI moderated the associations between PND and study outcomes. Analyses were performed in the whole study sample and within the PND dyads respectively to investigate to what extent PND and SSRI treatment influenced maternal and child outcomes.

## Results

### Prenatal and antenatal factors predicting PND status and SSRI treatment

Descriptive statistics for prenatal and antenatal factors are presented in Table 1.1 and Table 1.2. Pairwise comparison showed that compared with non-PND women, women with PND were younger, had given birth to more children (i.e., higher parity), had lower educational levels and lower income (Table 1.1). They also reported more alcohol and tobacco use during pregnancy, elevated levels of depression and anxiety before childbirth, and a higher proportion reported a lifetime history of depression diagnoses. Multivariable logistic regression including all the variables showed that lower education level (OR 0.86, 95%CI 0.83,0.89), lower income (OR 0.89, 95%CI 0.86,0.92), tobacco use during pregnancy (OR 1.13, 95%CI 1.02,1.26), lifetime history of depression (OR 2.06, 95%CI 1.88,2.21) and higher levels of antenatal depression and anxiety (OR 2.35, 95%CI 2.29,2.42) differentiated PND women from non-PND controls. Several prenatal and antenatal factors also differentiated SSRI-treated from non-SSRI-treated PND women (Table 1.2) including a lower parity (OR 0.74, 95%CI 0.59,0.92), a lower educational level (OR 0.84, 95%CI 0.71,0.99), a higher level of antenatal depression and anxiety (OR 1.16, 95%CI 1.05,1.28) and a higher proportion with a lifetime history of depression (OR 6.43, 95%CI 4.51,9.22). The postpartum EPDS scores were also significantly higher among the SSRI-treated PND group than the non-SSRI-treated group (OR 1.53, 95%CI 1.24,1.86).

**Table 1.1.** Antenatal maternal and child characteristics of non PND and PND dyads

|  | Non PND<br>n= 52038 | PND<br>N=8616 |  |  |
| --- | --- | --- | --- | --- |
|  | mean (sd) | mean (sd) | OR | Adj OR (95% CI) |
| **Maternal prenatal characteristics** |  |  |  |  |
| Maternal age | 30.41 (4.31) | 29.93 (4.76) | 0.98 (0.97, 0.98) *** | 1.00 (1.00, 1.01) |
| Parity | 0.74 (0.85) | 0.77 (0.87) | 1.05 (1.02, 1.08) *** | 1.01 (0.97, 1.05) |
| Maternal education |  |  |  |  |
| Ranking (scale 1-6) | 4.74 (1.16) | 4.38 (1.35) | 0.75 (0.73, 0.77) *** | 0.86 (0.83, 0.89) *** |
| Maternal Income |  |  |  |  |
| Ranking (scale 1-7) | 4.23 (1.32) | 3.87 (1.36) | 0.76 (0.74, 0.78) *** | 0.89 (0.86, 0.92) *** |
| Antenatal alcohol exposure |  |  |  |  |
| Ever used in pregnancy, N (%) | 14345 (27.6) | 2456 (28.5) | 1.10 (1.05, 1.16) *** | 1.05 (0.99, 1.12) |
| Antenatal tobacco exposure |  |  |  |  |
| Ever used in pregnancy, N (%) | 2494 (4.8) | 767 (8.9) | 1.99 (1.83, 2.16) *** | 1.13 (1.02, 1.26) * |
| Lifetime depression diagnosis, N (%) | 2230 (4.3) | 1381 (16.0) | 4.26 (3.96, 4.57) *** | 2.06 (1.88, 2.21) *** |
| **Antenatal maternal depression & anxiety (Gestational week 30)** |  |  |  |  |
| Scl-8 | 9.58 (2.13) | 12.48 (3.98) | 2.52 (2.46, 2.58) *** | 2.35 (2.29, 2.42) *** |
| **Postnatal depression (Postnatal month 6)** |  |  |  |  |
| EPDS | 1.98 (1.93) | 9.06 (2.17) | - | - |
| **Child characteristics** |  |  |  |  |
| Child sex (male) n (%) | 26558 (51) | 4441 (51.5) | - | - |
| Birth weight (kg) | 3.59 (0.55) | 3.58 (0.58) | 0.98 (0.96, 1.01) | - |
| Gestational age (weeks) | 39.74 (2.32) | 39.68 (2.43) | 0.97 (0.95, 1.00) * | - |

- a. Adjusted multiple logistic regression controlling for all the prenatal and antenatal factors to identify predictors for PND: reference group is non-PND dyads - b. Reported p value were adjusted for multiple comparison ( $FDR < 0.5$ ), \*\*\* $p < 0.001$ , \*\* $p < 0.01$ , \* $p < 0.05$

**Table 1.2.** Antenatal maternal and child characteristics of non-SSRI-treated PND and SSRI-treated PND

|  | Non-treated PND dyad<br>n=8440 | Treated PND dyad<br>n=176 | Treated<br>Vs.<br>Non- treated |  |
| --- | --- | --- | --- | --- |
|  | mean (sd) | mean (sd) | OR (95% CI) | Adj OR (95% CI) |
| **Maternal prenatal characteristics** |  |  |  |  |
| Maternal age | 29.94 (4.76) | 30.20 (5.01) | 1.01 (0.98, 1.04) | 1.04 (1.00, 1.09) |
| Parity | 0.78 (0.87) | 0.68 (0.84) | 0.88 (0.73,1.05) | 0.74 (0.59, 0.92) * |
| Maternal education |  |  |  |  |
| Ranking (scale 1-6) | 4.39 (1.35) | 4.14 (1.42) | 0.85 (0.74,0.97) * | 0.84 (0.71, 0.99) * |
| Maternal Income |  |  |  |  |
| Ranking (scale 1-7) | 3.88 (1.36) | 3.71 (1.39) | 0.88 (0.76, 1.03) | 0.98 (0.81, 1.19) |
| Antenatal alcohol exposure |  |  |  |  |
| ever used in pregnancy, N (%) | 2408 (28.5) | 48 (27.3) | 0.92 (0.65, 1.29) | 0.86 (0.60,1.23) |
| Antenatal tobacco exposure |  |  |  |  |
| Ever used in pregnancy, N (%) | 742 (8.8) | 25 (14.2) | 1.68 (1.07, 2.54) * | 0.93 (0.53, 1.54) |
| Lifetime depression diagnosis, N (%) | 1276 (15.1) | 105 (59.7) | 8.30 (6.12, 11.32) *** | 6.43 (4.51, 9.22) *** |
| **Antenatal maternal depression & anxiety (Gestational week 30)** |  |  |  |  |
| SCL-8 | 12.42 (3.92) | 15.29 (5.43) | 1.41 (1.31, 1.52) *** | 1.16 (1.05, 1.28) *** |
| **Postnatal depression (Postnatal month 6)** |  |  |  |  |
| EPDS | 9.03 (2.15) | 10.57 (2.63) | 2.09 (1.77, 2.44) *** | 1.53 (1.24, 1.86) *** |
| **Child characteristics** |  |  |  |  |
| Child sex (male) n (%) |  |  |  |  |
| Birth weight (kg) | 3.59 (0.57) | 3.53 (0.53) | - | - |
| Gestational age (weeks) | 39.71 (2.42) | 39.32 (2.14) | - | - |

- Adjusted multiple logistic regression controlling for all the prenatal and antenatal factors to identify predictors for SSRI treatment: reference group is non-treated PND dyads - Reported p value were adjusted for multiple comparison (FDR< 0.5), \*\*\*p<0.001, \*\* p<0.01, \*p<0.05

Prenatal and antenatal factors differentiating SSRI treatment status were used to generate a propensity score. The included factors were parity, maternal education, antenatal maternal depression and anxiety and lifetime depression history.

### The association between PND, SSRI treatment and maternal and child outcomes in non-PND dyads and PND dyads

Maternal and child outcomes in non-PND dyads and PND dyads are shown in *Table 2*. Table 2 also presents crude beta estimates from univariable regressions using SCL-8 at postpartum month 6 as the predictor variable and estimates from multiple regressions with covariates and SCL-8*SSRI moderation terms. Univariable analyses showed that PND symptoms were associated with higher levels of depression and anxiety across postpartum year 1.5 to year 5, and poorer relationship satisfaction across postpartum month 6 to year 3. PND symptoms were associated with higher levels of child internalizing and externalizing behaviors measured across ages 1.5 to 5 years, elevated ADHD symptoms at age 5 and poorer motor and language development at years 1.5 and 3. The associations remained significant, if somewhat attenuated, after adjustment for covariates.

**Table 2.1.**
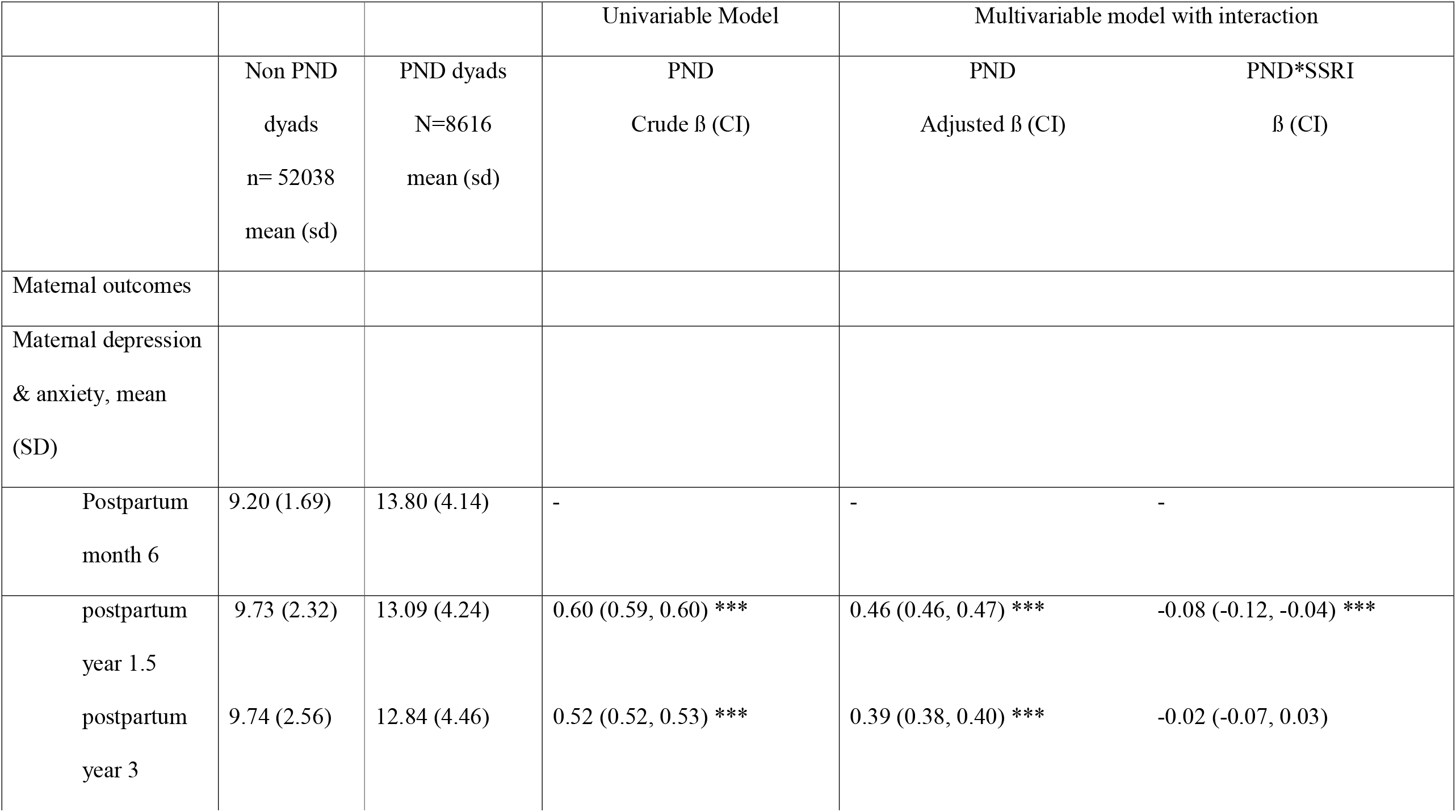
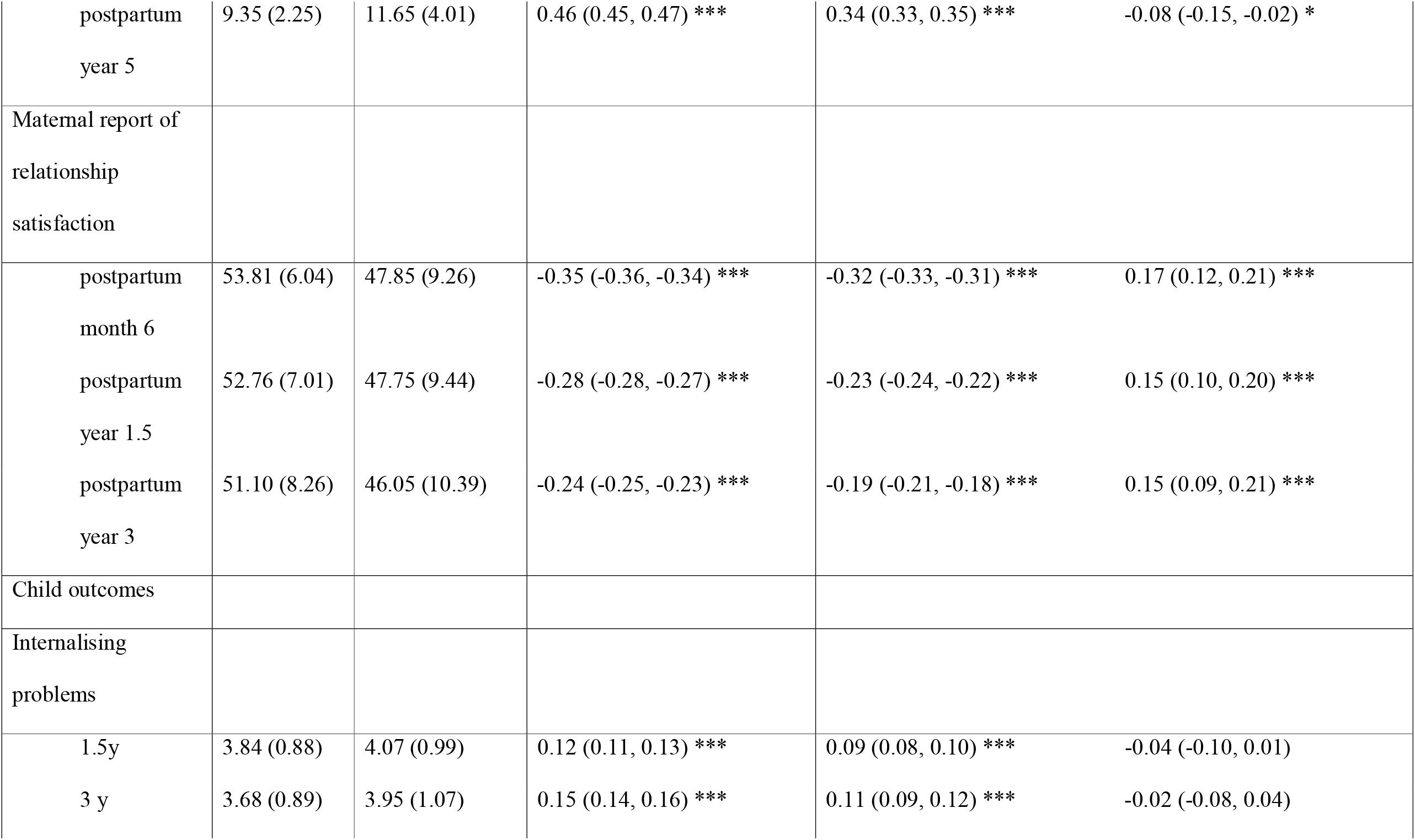
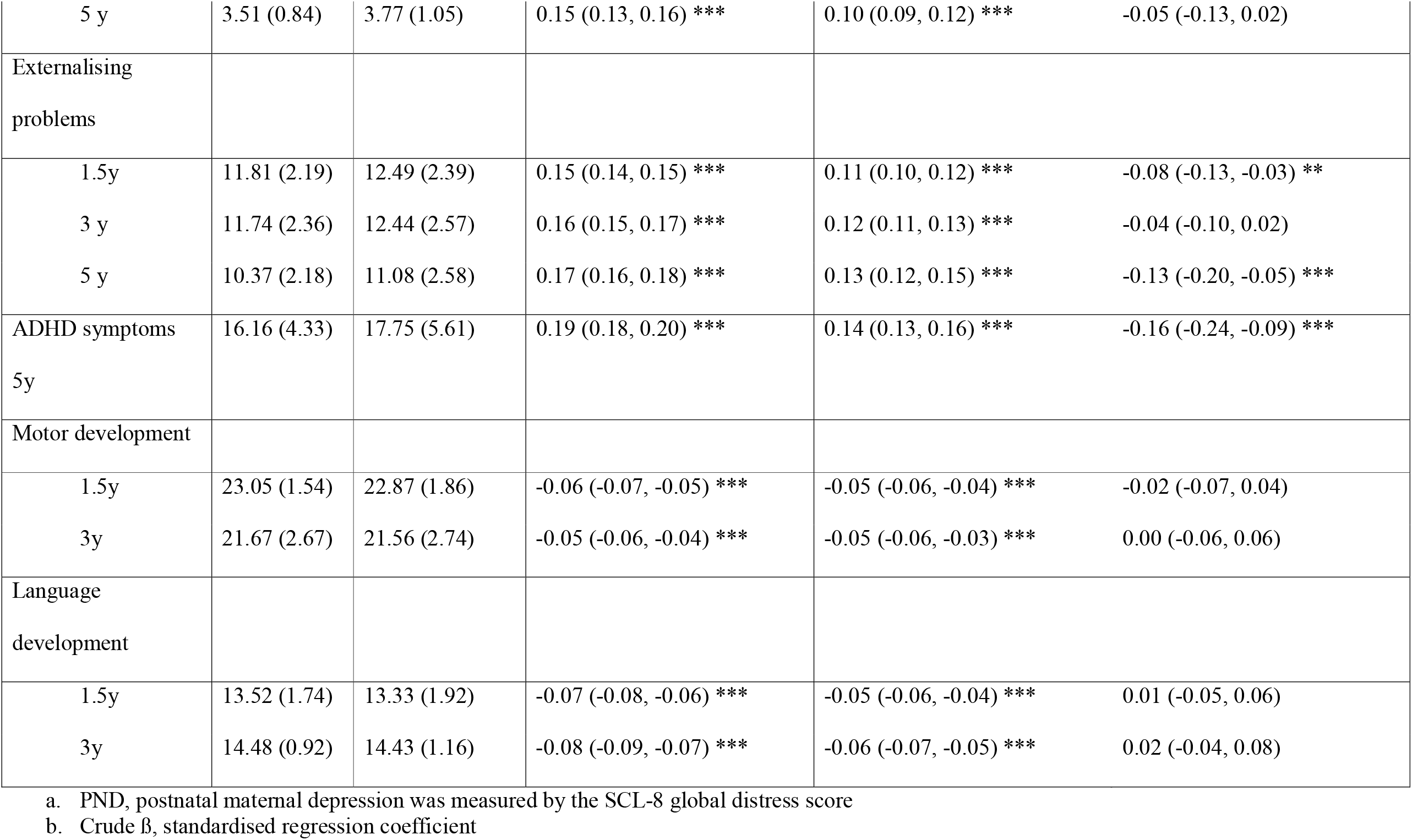
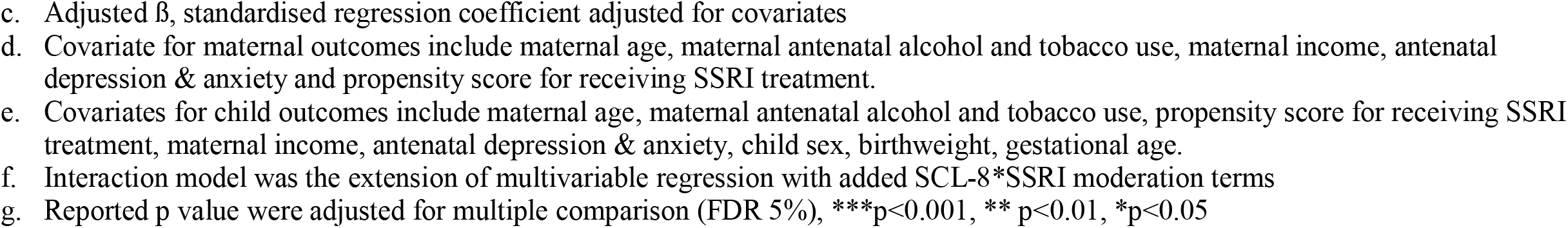
Maternal and child outcomes and the association between postnatal maternal depression, SSRI treatment and the study outcomes

**Table 2.2.**
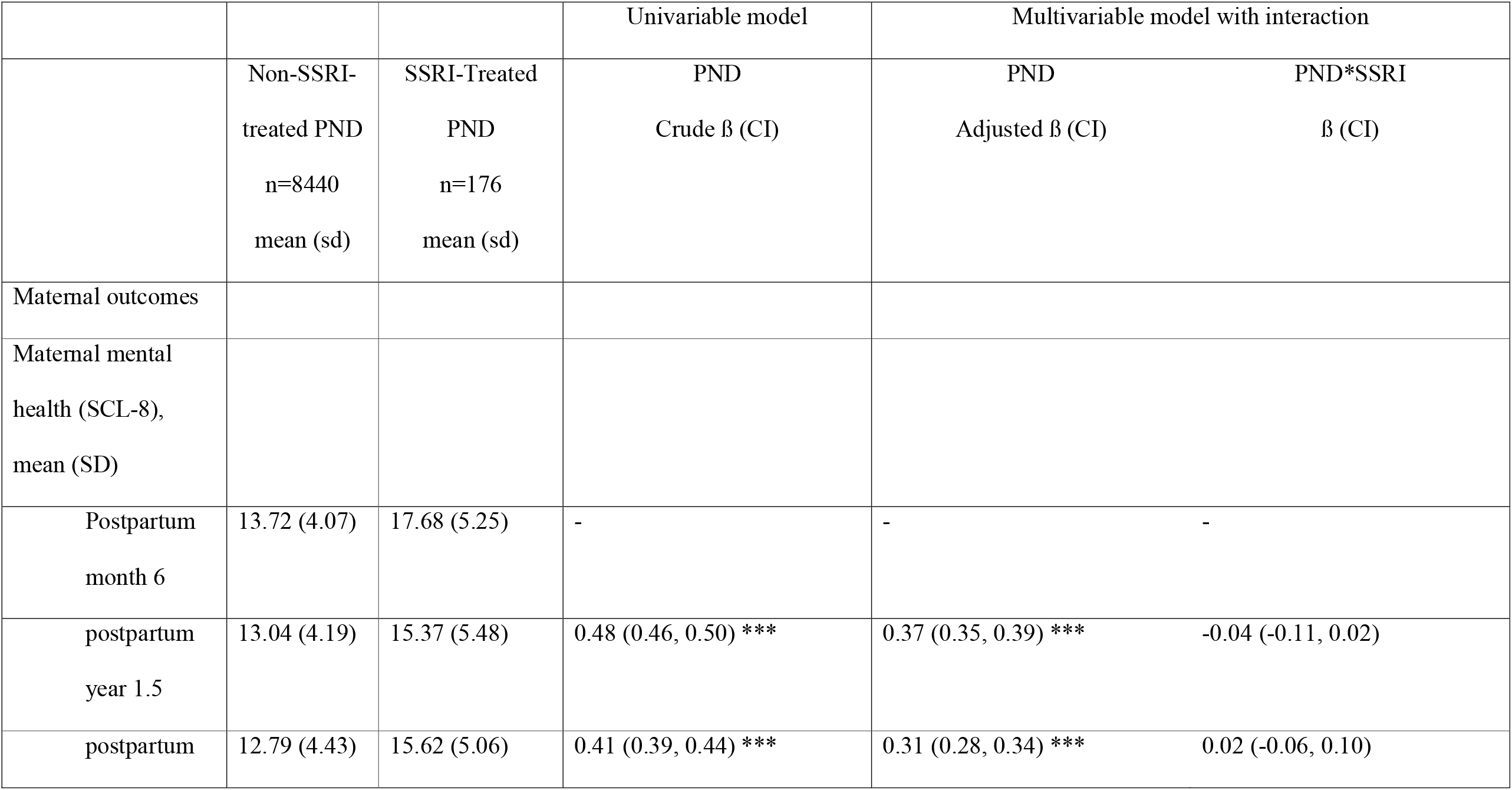
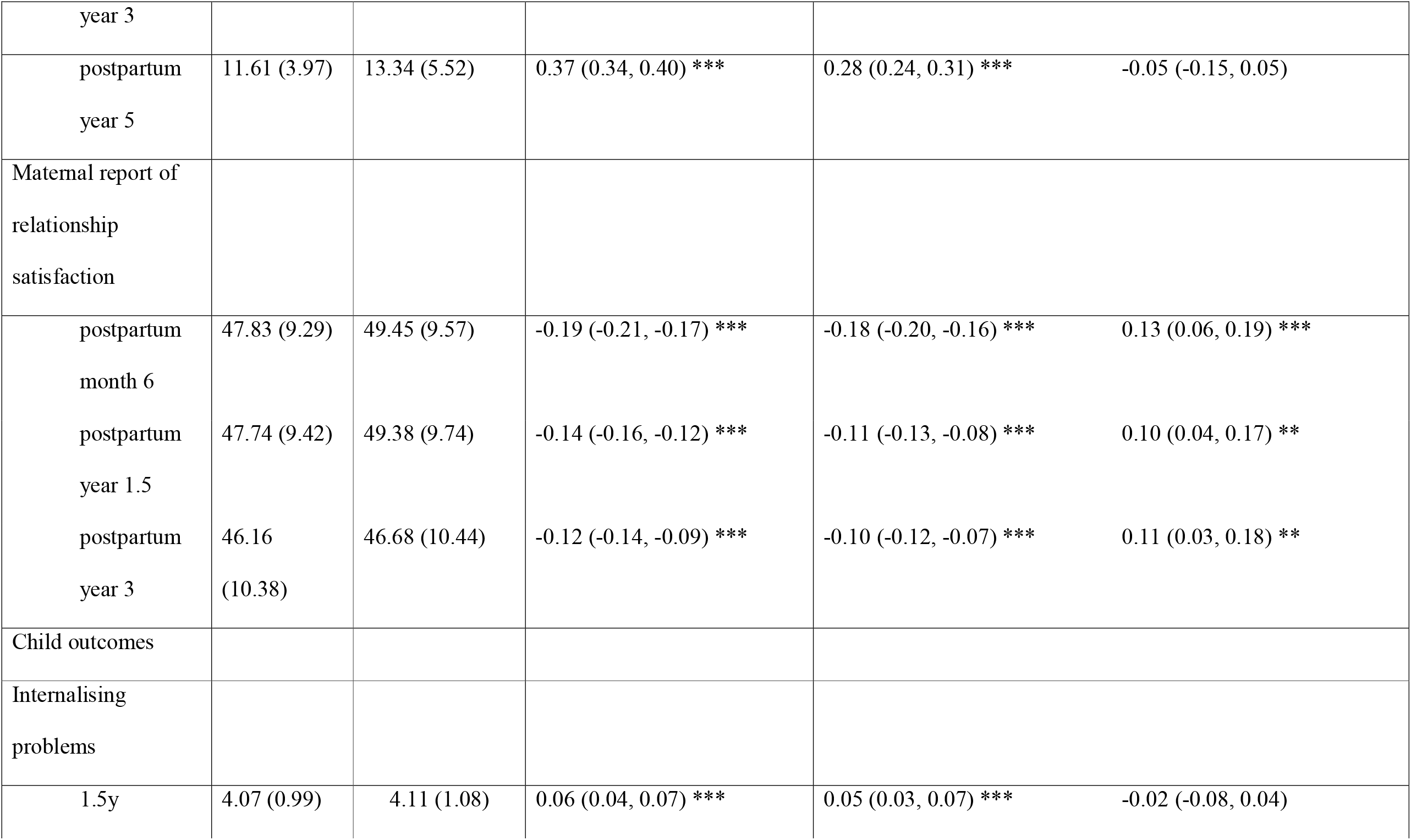
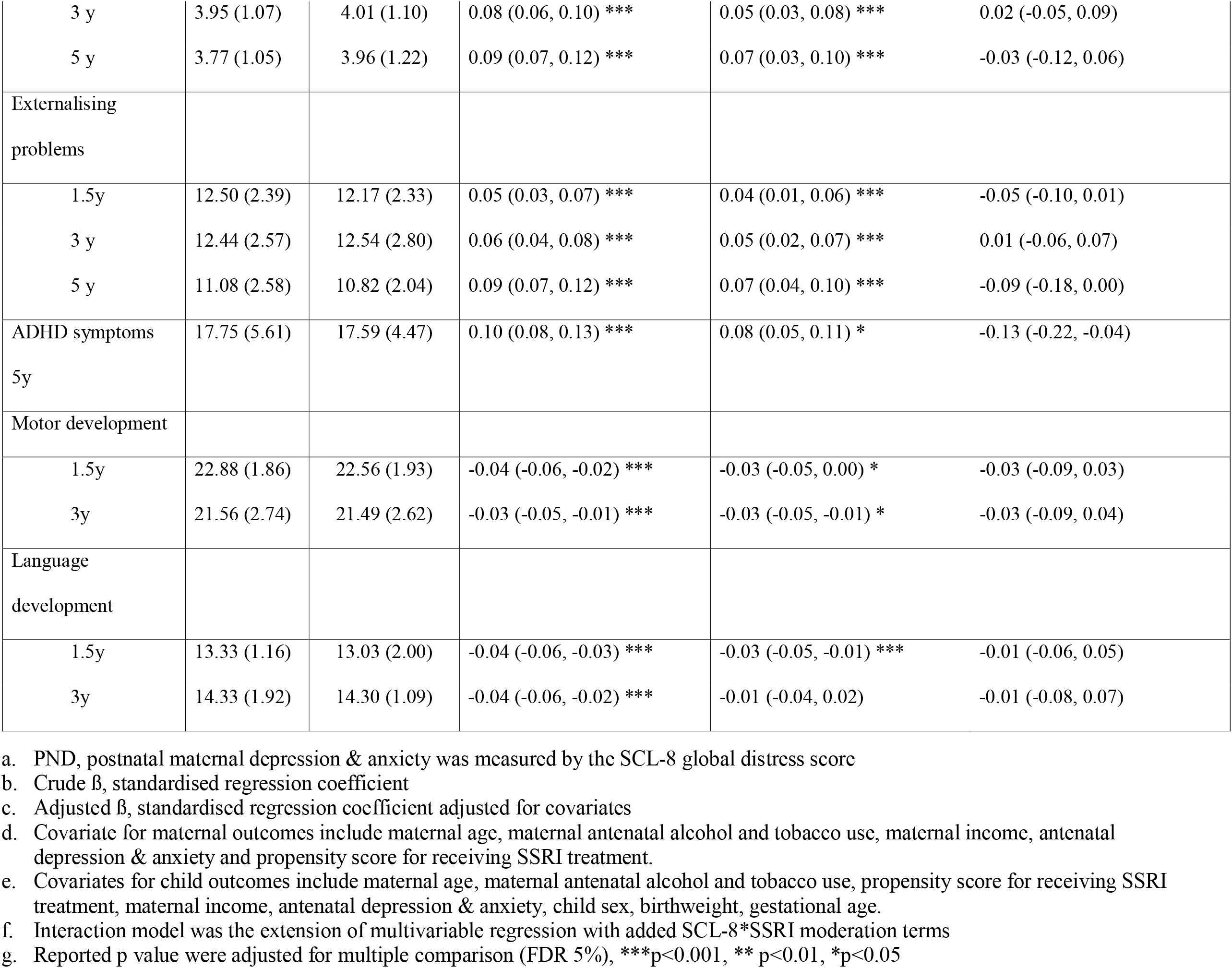
Maternal and child outcomes and the association between postnatal maternal depression, SSRI treatment and the study outcomes between treated and non-treated PND

### Moderation effects of SSRI treatment on the relationships between PND and maternal and child outcomes

Table 2.1 presents associations between PND symptoms, SSRI treatment and study outcomes in the whole study sample. SSRI treatment significantly reduced the negative impact of PND on the following maternal outcomes: maternal depression at postpartum year 1.5 and postpartum year 5 and relationship satisfaction at postpartum month 6, years 1.5 and 3. SSRI treatment also significantly reduced the associations between PND and the following child outcomes: externalizing behaviors at ages 1.5 and 5 years, and ADHD symptoms at age 5 years. There was no evidence that SSRI treatment moderated the associations between PND and child internalizing behaviors, or motor and language development across the 5-year follow up. The direction of effects for all interaction terms (significant and non-significant) except for child language development indicated that maternal SSRI use *did not or was very unlikely to* worsen associations between postnatal depression and any of our outcomes. The associations between postnatal depression, SSRI treatment and study outcomes among PND dyads are shown in Table 2.2. Evidence supports that SSRI treatment significantly attenuated the negative associations between PND and maternal relationship satisfaction. The direction of effects for other interaction terms converged with the findings in the whole study sample, indicating that SSRI use was *very unlikely* to worsen associations between postnatal depression and maternal mental health and child developmental outcomes.

## Discussion

We used data from a population-based cohort to study long-term outcomes for mother-child dyads affected by postnatal depression (PND) and provide insights into whether SSRI use moderated any observed relationships. We found that PND was associated with poorer maternal and child outcomes up to 5 years postpartum. Women with PND who received SSRIs presented with more severe postnatal depression than their non-SSRI-treated counterparts and were more likely to have a prior history of depression. Overall, our analyses indicated that treatment with SSRIs attenuated the negative associations between PND and subsequent maternal depression and child externalizing behaviors. We found no evidence to suggest that SSRIs conferred an increased risk for any of the study outcomes.

### Prenatal and antenatal factors differentiated PND and the use of SSRI

When multiple prenatal and antenatal factors were examined together, higher levels of antenatal depression and anxiety and a lifetime depression diagnosis were associated with the highest risk for PND. These findings are in line with meta-synthesised data indicating that depression and anxiety during pregnancy and a previous history of depression conferred the largest risks of PND, greater than factors such as pregnancy related complications, maternal neuroticism, and socioeconomic adversity [42]. It is noteworthy that the strong risk (2 times increased) associated with a lifetime diagnosis of depression in the occurrence of PND was observed independently of the levels of antenatal depression and anxiety in the current study. Such findings are consistent with the knowledge that recurrent mood episodes during the postpartum period are common in women with pre-existing major depression [43]. More importantly, it suggests that prenatal screening for maternal mental health history can be useful in signposting a heightened risk of PND.

Interestingly, although postnatal depression levels were significantly different between the SSRI-treated and non-SSRI-treated women, a previous history of depression independently predicted the likelihood of SSRI use (OR 6.43 for a history of depression vs. OR 1.53 for EPDS level). In addition, the use of SSRI was predicted by lower parity and lower maternal education. This suggests that PND severity was not the sole determinant of the use of SSRIs in our sample. Indeed, information gathered from health professionals and women with PND suggests that awareness of mental health problems and socioeconomic status predicted antidepressant use for postnatal depressive symptoms [23, 44, 45]. For example, postnatal depressive symptoms is at times viewed as maladjustment to motherhood by health providers and affected women, and hence medical treatment not considered [45]. Although reflection on the mechanism that leads to the use of SSRIs for PND remains speculative, our findings suggest that women with a history of depression are more open to the idea of pharmacological intervention perhaps due to better awareness of their mental health conditions. Such views have been documented among women with PND in prior research [22, 46].

### PND and maternal and child outcomes

In our cohort, PND symptoms predict elevated levels of postnatal maternal depression, anxiety, and relationship dissatisfaction up until 5 years after childbirth. Postnatal maternal anxiety has been associated with a range of negative developmental effects in children, including impaired socio-emotional development and increased externalizing behaviours [47, 48]. Partner relationship dissatisfaction is an established risk factor for marital conflict, increased psychological distress among family members, and poor parental involvement in childcare [7, 49]. It is noteworthy that relationship dissatisfaction also predicts the increase and persistence of PND symptoms [50]. These findings echo previous studies in suggesting that harmful consequences associated with PND can be found not only in affected women but also in the wider family [5, 12]. Our findings on the associations between PND and child outcomes are in line with related research which shows that children of mothers with PND are at risk of behavioral problems and developmental delay [25]. Crucially, our study and others have identified links between PND and long-term impairment of cognitive, emotional and behavioral functioning in exposed children [13, 51, 52]. The findings highlight the importance of early intervention for PND to prevent unfavorable outcomes in offspring in the long term.

### Moderation of SSRI treatment on the relationship between PND and postnatal outcomes

Considering the negative associations PND has across a wide range of outcomes, adequate treatment for PND is crucial. Although meta-analyses of randomised control trials have identified no adverse effects associated with SSRIs on breastfed infants, the limited evidence on the long-term consequences in children has led to treatment hesitancy in women with PND [14, 22]. The dilemma of whether depressive symptoms during the postnatal period should be regarded as a mental disorder, combined with uncertainty regarding the long-term safety of postnatal antidepressant use, also affect clinical decision and management approaches among health professionals [53]. The current study used longitudinal data on a large mother-child cohort to investigate benefits and risks associated with SSRI treatment for PND. We found that SSRI treatment mitigated the effects of PND on subsequent maternal depression and relationship dissatisfaction. That the positive effects of SSRI treatment on postpartum maternal mental health were observed up to 5 years after childbirth further indicates that SSRI treatment reduces the risks of depression persistence and is beneficial to the family environment in the long term. Our study also identified that SSRI use mitigated the negative associations between PND and child outcomes. Specifically, our results suggest that SSRI treatment can effectively reduce the negative associations between PND and externalizing behaviors and ADHD symptoms at age 5 years. These findings support the notion that adequate treatment for PND may normalize child behavioral problems associated with the condition [54]. With regard to the concern of long-term safety for children, we found no evidence that SSRIs conferred an increased risk of problems in motor and language development up to 5 years of age.

## Limitations

A major strength of our study is the longitudinal design with repeated measurement in a large mother-child cohort. Our findings fill a gap in the literature in providing data on long-term child outcomes associated with SSRI use for women with PND. As hypothesized, evidence shows that SSRI treatment brings benefits to affected women and their family in the long term. Our study did not identify additional risks associated with postnatal SSRI use in child emotional, behavioral, motor and language development.

Notwithstanding the strengths, several limitations pertinent to the study design warrant mention. First, the study used maternal self-report data from a population-based cohort. Previous research indicates that depressed mothers tended to over-report emotional and behavioral problems of their children and the reporting errors tend to increase with the severity of depression [55, 56]. SSRI-treated women in our sample presented with more severe depression throughout the perinatal period, and therefore they may systematically over-report problematic symptoms in their children. However, it also suggests that the positive effects associated with SSRIs on depressed mothers and their children are less likely to result from depression-related reporting bias. Future studies should incorporate different sources of reports to evaluate this issue. Second, the current study used self-reported medication history for SSRI use. Although self-report medication use may be a potentially inaccurate, the concordance between self-reports and records from the national prescription database for perinatal SSRI use is high in the current study sample (MoBa) [57]. Third, plasma concentrations of antidepressant in infants were not available in MoBa due to the observational nature of the study design. Although clinical studies have found different plasma concentrations in breastfed infants for different SSRIs, for SSRIs recommended as the first-line treatment for PND such as sertraline and paroxetine, the infant plasma concentrations are very low or undetectable [21]. Indeed, to better understand pharmacological effects associated with the use of SSRIs, information on infant plasma concentration would be ideal to control for exposure level. However, issues of feasibility would likely arise in any attempts to perform timed and repeated blood tests for infants exposed to SSRI in a large longitudinal sample such as the one we analysed. Another limitation regarding data availability is that we did not know whether women received non-pharmacological intervention alongside (or instead of) SSRIs. The effects associated with SSRIs may be exaggerated if the women simultaneously received other forms of treatment for PND or weakened if women in the ‘non-SSRI-treated’ dyad presented with better outcomes due to other interventions. We acknowledge the aforementioned limitations pertinent to observational study design and our findings should be interpreted with caution.

However, given the costs and ethical concerns that would be associated with conducting large-scale randomised control trials for the long-term consequences associated with SSRI use in the management for PND, we suggest that our findings represent an important step forward in understanding.

## Conclusion

In this longitudinal study we aimed to increase understanding of the long-term outcomes associated with SSRI treatment of mother-child dyads affected by PND. Postnatal depression was associated with poorer maternal and child mental health outcomes and child developmental outcomes up to postpartum year 5. However, the associations between PND and some unfavourable outcomes were attenuated by SSRI treatment. We did not find evidence that postnatal SSRI exposure increased the risk of child’s emotional and behavioral problems or delayed motor and language development. While our findings need to be replicated, we believe that they add important information regarding long-term outcomes associated with SSRI use for postnatal maternal depression. Such information might help affected women and their family to better understand the benefits and risks of taking SSRIs and make informed decisions regarding PND treatment.

## Data Availability

All data produced are available upon request at https://www.fhi.no/en/studies/moba/for-forskere-artikler/research-and-data-access/

## Acknowledgements

The Norwegian Mother, Father and Child Cohort Study is supported by the Norwegian Ministry of Health and Care Services and the Ministry of Education and Research. We are grateful to all the participating families in Norway who take part in this on-going cohort study. The positions of CYL and TAM are supported by a Wellcome Trust Senior Research Fellowship awarded to TAM [220382/Z/20/Z]. EY was supported by the Norwegian Research Council (262177 and 288083). **For the purpose of open access, the author has applied a CC BY public copyright licence to any Author Accepted Manuscript version arising from this submission**

---

**Supplementary Table 1.** Outcome of treated and non-treated PND

|  | Non-treated PND dyads<br>n=8440 mean (sd) | Treated PND dyads<br>n=176<br>mean (sd) | Effect sizes |
| --- | --- | --- | --- |
| Maternal outcomes |  |  |  |
| Maternal mental health (SCL-8), mean (SD) |  |  |  |
| postpartum month 6 | 13.72 (4.07) | 17.68 (5.25) | 0.11 *** |
| postpartum year 1.5 | 13.04 (4.19) | 15.37 (5.48) | 0.06 *** |
| postpartum year 3 | 12.79 (4.43) | 15.62 (5.06) | 0.07 *** |
| postpartum year 5 | 11.61 (3.97) | 13.34 (5.52) | 0.03 ** |
| Maternal depression, mean (SD) | 13.72 (4.07) | 17.68 (5.25) |  |
| postpartum month 6 | 7.45 (2.41) | 9.28 (3.06) | 0.09 *** |
| postpartum year 1.5 | 7.07 (2.47) | 8.17 (3.35) | 0.04 *** |
| postpartum year 3 | 6.90 (2.62) | 8.23 (3.05) | 0.05 *** |
| postpartum year 5 | 6.20 (2.37) | 7.01 (3.34) | 0.02 |
| Maternal anxiety, mean (SD) |  |  |  |
| postpartum month 6 | 6.27 (2.15) | 8.42 (2.81) | 0.12 *** |
| postpartum year 1.5 | 5.97 (2.12) | 7.21 (2.65) | 0.07 *** |
| postpartum year 3 | 5.89 (2.17) | 7.39 (2.45) | 0.08 *** |
| postpartum year 5 | 5.43 (1.92) | 6.32 (2.51) | 0.04 *** |
| Maternal report of relationship satisfaction |  |  |  |
| postpartum month 6 | 47.83 (9.29) | 49.45 (9.57) | 0.03 * |
| postpartum year 1.5 | 47.74 (9.42) | 49.38 (9.74) | 0.03 * |
| postpartum year 3 | 46.16 (10.38) | 46.68 (10.44) | 0.01 |
| Child outcomes |  |  |  |
| Internalising problems |  |  |  |
| 1.5y | 4.07 (0.99) | 4.11 (1.08) | 0.00 |
| 3 y | 3.95 (1.07) | 4.01 (1.10) | 0.01 |
| 5 y | 3.77 (1.05) | 3.96 (1.22) | 0.02 |
| Externalising problems |  |  |  |
| 1.5y | 12.50 (2.39) | 12.17 (2.33) | 0.02 |
| 3 y | 12.44 (2.57) | 12.54 (2.80) | 0.00 |
| 5 y | 11.08 (2.58) | 10.82 (2.04) | 0.00 |
| ADHD symptoms 5y | 17.75 (5.61) | 17.59 (4.47) | 0.01 |
| Motor development |  |  |  |
| 1.5y | 22.88 (1.86) | 22.56 (1.93) | 0.03 |
| 3y | 21.56 (2.74) | 21.49 (2.62) | 0.01 |
| Language development |  |  |  |
| 1.5y | 13.33 (1.16) | 13.03 (2.00) | 0.02 |
| 3y | 14.33 (1.92) | 14.30 (1.09) | 0.01 |

a. Reported p value were adjusted for multiple comparison (FDR 5%), \*\*\*p<0.001, \*\* p<0.01, \*p<0.05

